# Common and distinct cortico-striatal volumetric changes in cocaine and heroin use disorder

**DOI:** 10.1101/2022.02.27.22271574

**Authors:** Ahmet O Ceceli, Yuefeng Huang, Greg Kronberg, Pias Malaker, Pazia Miller, Sarah King, Pierre-Olivier Gaudreault, Natalie McClain, Lily Gabay, Devarshi Vasa, Defne Ekin, Nelly Alia-Klein, Rita Z Goldstein

## Abstract

Drugs of abuse impact cortico-striatal dopaminergic targets and their morphology across substance types in common and unique ways. While the dorsal striatum drives addiction severity across drug classes, opiates impact ventromedial prefrontal cortex (vmPFC) and nucleus accumbens (NAcc) neuroplasticity in preclinical models, and psychostimulants alter inhibitory control, rooted in cortical regions such as the inferior frontal gyrus (IFG). We hypothesized parallel gray matter volume (GMV) changes in individuals with cocaine or heroin use disorder (CUD/HUD): decreased GMV of vmPFC/NAcc in HUD and IFG in CUD, and putamen GMV to be associated with addiction severity. We quantified GMV in age/sex/IQ-matched individuals with CUD (n=20; 5 women), HUD (n=20; 6 women), and healthy controls (HC; n=20; 5 women), further replicated in an extended sample (combined n=96). Overall, addicted individuals had smaller vmPFC volumes than HC (*p*<0.05-corrected), driven by HUD (*p*<0.05-corrected; similar NAcc reduction). Right IFG reductions were specifically evident in CUD vs. HUD (*p*<0.05-corrected). Posterior putamen volume increased as a function of craving in CUD vs. HUD (p<0.05-corrected). These results indicate compression of dopamine-innervated regions (in the vmPFC and NAcc) across cocaine- or heroin-addicted individuals, more severely in the latter. For the first time we demonstrate IFG compression specifically in CUD. This group also showed a unique association between craving and increased putamen volume, together indicating a signature of enhanced cue-sensitivity and habit formation. Results suggest common and substance-specific morphometry volumetric changes in human psychostimulant or opiate addiction, with implications for fine-tuning biomarker and treatment identification by primary drug of abuse.

## Introduction

Psychostimulants and opiates are among two of the most treatment-resistant drugs of abuse (1), together contributing to over 100,000 deaths in the United States annually (2). Drugs of abuse converge in their dopamine receptor-targeted mechanisms of action (3), as associated with the morphology of the mesocorticolimbic regions that dopamine innervates (4). Drugs of abuse also demonstrate distinct neurobehavioral signatures (5), together suggesting both common and unique effects on behavior and brain volume. For example, volumetric decreases, exacerbated with addiction severity (e.g., years of use), have been reported in drug-addicted individuals for the ventromedial prefrontal cortex (vmPFC) (6–9). These vmPFC volumetric decreases potentially contribute to deficits in this region’s core functions, namely salience/value processing (10), extinction learning (11), and goal-directed control (12), a cognitive and emotional pattern contributing to the drug addiction phenomenology (13,14).. In an opposite direction but still common to drug-addicted individuals (i.e., spanning different drug classes), volumetric increases have been reported for the dorsal striatum (15) that regulates habit formation (16,17), potentially related to its dopaminergically-mediated role in cue sensitivity and craving (18).

On the other hand, chronic self-administration of heroin, but not cocaine (19), in rats contributes to decreased dendritic spine density in the vmPFC/orbitofrontal cortex (OFC) (20,21) and impaired extinction learning (e.g., persistent drug seeking despite unavailability of the drug) (22), suggesting an especially robust impact of heroin on the vmPFC. The nucleus accumbens (NAcc), a motivational hub with core roles in perceived salience (23,24), reward anticipation (25), and drug craving (23), has also been a major focus of investigation in these studies. Similarly to the direction of impact on the vmPFC/OFC, chronic opiate self-administration in rats is associated with decreased measures of neuroplasticity (dendritic branching and spine density) in the medium spiny neurons of the NAcc (21), a result in line with NAcc gray matter volume (GMV) reductions in human heroin addiction (26). In contrast, escalation to cocaine but not heroin self-administration is more likely in rats that are high in trait impulsivity (27,28). Similarly in humans, impulsivity and inhibitory control deficits, regulated by cortical regions including the inferior frontal gyrus (IFG) (29–31), may be more closely aligned with the symptom profile of cocaine addiction (5,32). Consistent with this suggestion, a study of 169 male polysubstance users (cocaine: ≥1 gram/week; alcohol: ≥21 drinks/week for heavy, 1-21 drinks/week for light drinkers; marijuana: ≥1 joint/week; tobacco: >1 cigarette/day) revealed decreased IFG volumes as a function of monthly cocaine use when controlling for the use of the other substances, although this study did not account for opiate use (33).

Complementing the mostly preclinical efforts, several meta-analyses of GMV studies in human addiction similarly allow the comparison between different drugs classes (6,7,9,34,35). These meta-analyses support shared vmPFC reductions across substances (6,7,9) and suggest several substance-specific patterns [e.g., alcohol-specific reductions in the NAcc (6), and cocaine-specific (relative to methamphetamine) reductions in the superior frontal gyrus (34)]. However, to the best of our knowledge, only two studies to date have directly assessed the overlapping and distinct neuroanatomical underpinnings related to psychostimulant and opiate use as they manifest in gray matter (36,37). In one study, group comparisons revealed GMV reductions in parietal and precentral/postcentral gyrus in 28 individuals with a history of heroin abuse, and in hippocampus, posterior cingulate cortex, and cerebellum in 14 individuals with a history of cocaine abuse (36). In the other study, compared to a group of 21 individuals with polysubstance use disorder that included cocaine and alcohol use, a group of 27 individuals with opioid use disorder (not specific to heroin) had cortical thinning in the posterior superior frontal gyrus and temporo-parietal junction (37). Notably, opposite to expectations from the preclinical literature, in these studies the vmPFC, NAcc, IFG or dorsal striatum were not implicated.

Here, to investigate the common and distinct cortico-striatal GMV patterns related to opiate or stimulant addiction, we used voxel-based morphometry (VBM) of T1-weighted magnetic resonance imaging (MRI) in demographically matched groups of individuals with cocaine or heroin use disorder (CUD/HUD) and healthy controls (HC), with an internal replication of analyses in an extended sample. We aimed to address the following hypotheses: 1) compared to HC, drug-addicted individuals would exhibit reduced GMV in the vmPFC, with this reduction most pronounced in HUD, 2) the HUD group would show reduced GMV in the NAcc, 3) the IFG would be smaller in CUD compared to HUD, and 4) the common and/or unique neuroanatomical patterns in the CUD and HUD would be further supported by potentially distinct associations with addiction severity measures including craving, a measure of cue sensitivity and habitual associations; here, we speculated a role for the dorsal striatum.

## Methods

### Participants

Three age-, sex-, education- and IQ-matched groups of 20 individuals with HUD (40.9±10 years, 6 women), 20 individuals with CUD (42.8±7 years, 5 women), and 20 HC (41.4±9 years, 5 women), comprised our main sample of 60 participants. An additional 12 participants per group yielded a larger total sample (n=96) for internal replication purposes (see Table 1 for details on both samples). Subjects with CUD were recruited by advertisements and flyers (in local newspapers, bulletin boards, and online) as well as from educational talks provided to groups of staff and patients at collaborating substance abuse prevention and treatment institutes in the New York metropolitan area. HC were recruited from the same communities for matching purposes. Individuals with HUD were recruited from a single inpatient drug addiction rehabilitation facility (Samaritan Daytop Village, NY). The Icahn School of Medicine at Mount Sinai’s institutional review board approved study procedures, and all participants provided written informed consent.

**Table 1.**
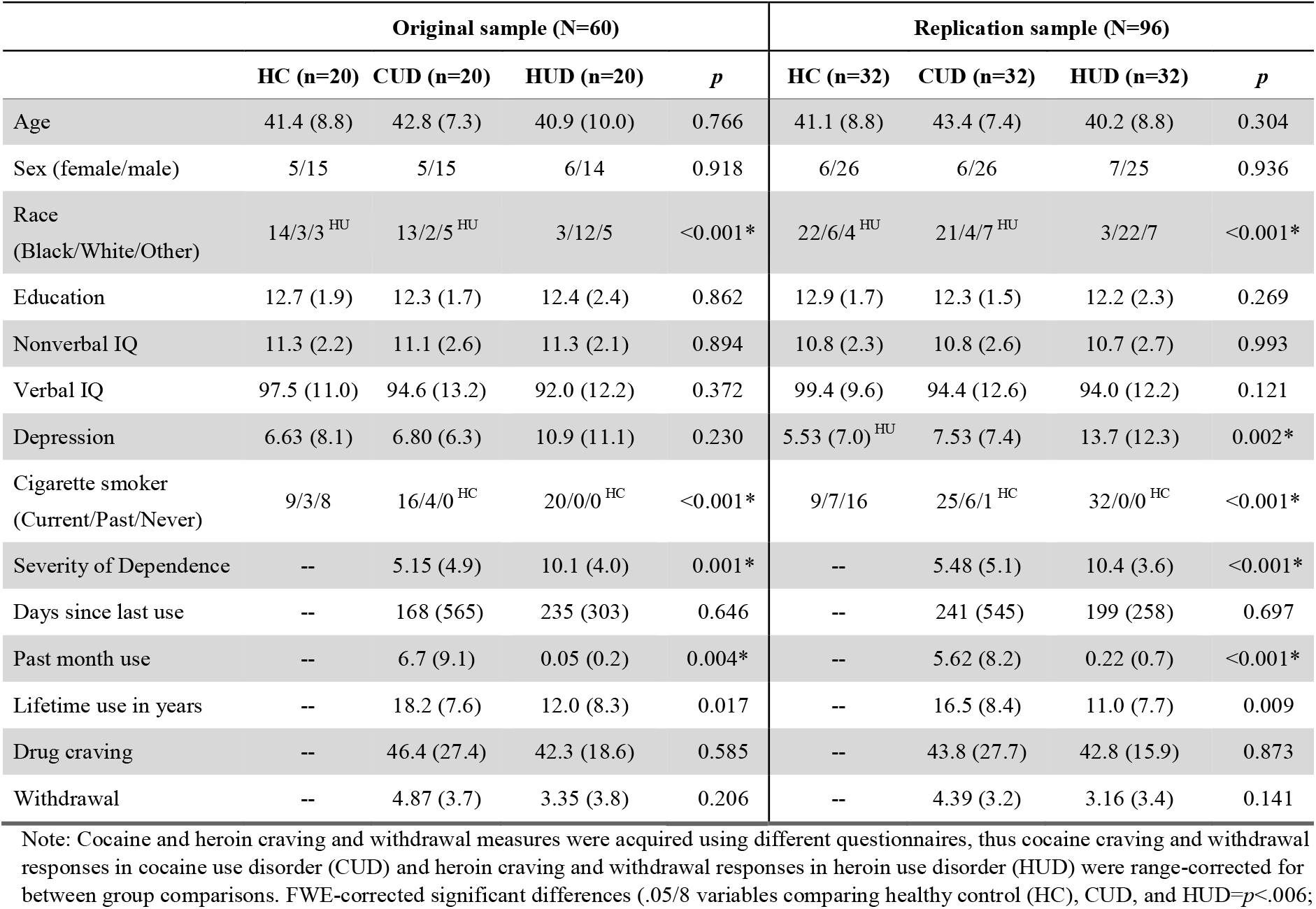
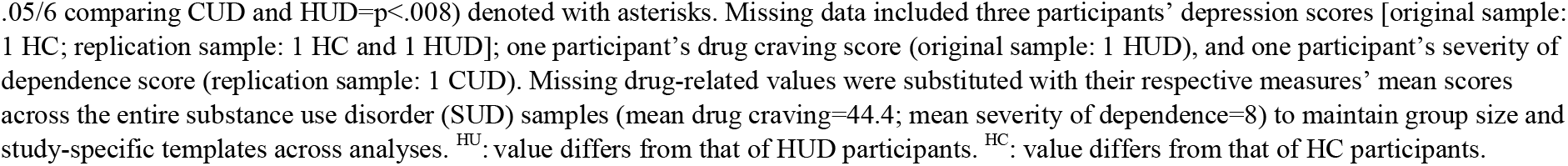
Sample profile.

| Original sample (N=60) |  |  |  |  | Replication sample (N=96) |  |  |  |
| --- | --- | --- | --- | --- | --- | --- | --- | --- |
|  | HC (n=20) | CUD (n=20) | HUD (n=20) | *p* | HC (n=32) | CUD (n=32) | HUD (n=32) | *p* |
| Age | 41.4 (8.8) | 42.8 (7.3) | 40.9 (10.0) | 0.766 | 41.1 (8.8) | 43.4 (7.4) | 40.2 (8.8) | 0.304 |
| Sex (female/male) | 5/15 | 5/15 | 6/14 | 0.918 | 6/26 | 6/26 | 7/25 | 0.936 |
| Race<br>(Black/White/Other) | 14/3/3<sup>HU</sup> | 13/2/5<sup>HU</sup> | 3/12/5 | <0.001* | 22/6/4<sup>HU</sup> | 21/4/7<sup>HU</sup> | 3/22/7 | <0.001* |
| Education | 12.7 (1.9) | 12.3 (1.7) | 12.4 (2.4) | 0.862 | 12.9 (1.7) | 12.3 (1.5) | 12.2 (2.3) | 0.269 |
| Nonverbal IQ | 11.3 (2.2) | 11.1 (2.6) | 11.3 (2.1) | 0.894 | 10.8 (2.3) | 10.8 (2.6) | 10.7 (2.7) | 0.993 |
| Verbal IQ | 97.5 (11.0) | 94.6 (13.2) | 92.0 (12.2) | 0.372 | 99.4 (9.6) | 94.4 (12.6) | 94.0 (12.2) | 0.121 |
| Depression | 6.63 (8.1) | 6.80 (6.3) | 10.9 (11.1) | 0.230 | 5.53 (7.0)<sup>HU</sup> | 7.53 (7.4) | 13.7 (12.3) | 0.002* |
| Cigarette smoker<br>(Current/Past/Never) | 9/3/8 | 16/4/0<sup>HC</sup> | 20/0/0<sup>HC</sup> | <0.001* | 9/7/16 | 25/6/1<sup>HC</sup> | 32/0/0<sup>HC</sup> | <0.001* |
| Severity of Dependence | -- | 5.15 (4.9) | 10.1 (4.0) | 0.001* | -- | 5.48 (5.1) | 10.4 (3.6) | <0.001* |
| Days since last use | -- | 168 (565) | 235 (303) | 0.646 | -- | 241 (545) | 199 (258) | 0.697 |
| Past month use | -- | 6.7 (9.1) | 0.05 (0.2) | 0.004* | -- | 5.62 (8.2) | 0.22 (0.7) | <0.001* |
| Lifetime use in years | -- | 18.2 (7.6) | 12.0 (8.3) | 0.017 | -- | 16.5 (8.4) | 11.0 (7.7) | 0.009 |
| Drug craving | -- | 46.4 (27.4) | 42.3 (18.6) | 0.585 | -- | 43.8 (27.7) | 42.8 (15.9) | 0.873 |
| Withdrawal | -- | 4.87 (3.7) | 3.35 (3.8) | 0.206 | -- | 4.39 (3.2) | 3.16 (3.4) | 0.141 |

Note: Cocaine and heroin craving and withdrawal measures were acquired using different questionnaires, thus cocaine craving and withdrawal responses in cocaine use disorder (CUD) and heroin craving and withdrawal responses in heroin use disorder (HUD) were range-corrected for between group comparisons. FWE-corrected significant differences (.05/8 variables comparing healthy control (HC), CUD, and HUD= $p < .006$ ;
.05/6 comparing CUD and HUD= $p<.008$ ) denoted with asterisks. Missing data included three participants' depression scores [original sample: 1 HC; replication sample: 1 HC and 1 HUD]; one participant's drug craving score (original sample: 1 HUD), and one participant's severity of dependence score (replication sample: 1 CUD). Missing drug-related values were substituted with their respective measures' mean scores across the entire substance use disorder (SUD) samples (mean drug craving=44.4; mean severity of dependence=8) to maintain group size and study-specific templates across analyses. <sup>HU</sup>: value differs from that of HUD participants. <sup>HC</sup>: value differs from that of HC participants.

Participants underwent a series of clinical and neuropsychological assessments delivered by trained research staff under a clinical psychologist’s supervision. These assessments included the Mini International Neuropsychiatric Interview (38) (for the HUD) or the Structured Clinical Interview for DSM-IV or DMS-5 Axis I and II Disorders (39) (for the CUD and HC); the Addiction Severity Index (40), a semi-structured interview that assesses history and severity of substance use problems for all individuals with substance use disorder (SUD); the Cocaine Selective Severity Assessment (41) or Short Opiate Withdrawal Scale (42) to evaluate abstinence/withdrawal symptomology related to cocaine or heroin, respectively (range-corrected to a common scale for group comparisons in withdrawal); the Severity of Dependence scale for all SUD participants (43); and the Cocaine Craving Questionnaire (44) or the Heroin Craving Questionnaire (range-corrected to a common scale for group comparisons in craving) (45). Participants underwent a brief physical examination including urine drug toxicology, breath carbon monoxide and alcohol measurements, and review of medical history.

All individuals with CUD and HUD met criteria for SUD with cocaine or heroin as the primary drug of choice, respectively. Other comorbidities included alcohol use disorder (CUD original sample n=7, extended n=11; HUD original n=2, extended n=3), marijuana use disorder (CUD original n=2, extended n=7; HUD original n=1, extended n=1), sedative use disorder (CUD extended n=1; HUD original n=3, extended n=4), phencyclidine use disorder (CUD extended n=1), meth/amphetamine use disorder (CUD extended n=1; HUD original n=1, extended n=2), major depressive disorder (HUD original n=2, extended n=3), post-traumatic stress disorder (CUD original n=1, extended n=1), and specific phobias (CUD original n=1, extended n=2). All SUD comorbidities were in partial or sustained remission at the time of study. Within the CUD sample, four participants in the original and eight in the extended sample also met criteria for HUD, and within the HUD, five participants in the original and eight in the extended sample also met criteria for CUD; however, these individuals’ primary drug of abuse was identified to be congruent with their assignment to their respective groups. Among the CUD group, urine screen results confirmed the presence of cocaine in 10 participants in the original and 14 participants in the extended samples. All participants in the HUD group were under medication assisted treatment, with urine toxicology positive for: methadone (original n=18; extended n=26), buprenorphine (original n=1; extended n=5), and methadone and buprenorphine (original n=1, extended n=1). Mean methadone doses were 103.8 ± 63.3 mg (original sample, 2 missing) or 99.4 ± 56.4 mg (extended, 2 missing); buprenorphine 12 mg (original, 1 missing) or 14.7 ± 8.3 mg (extended, 3 missing). The route of drug administration included smoking (CUD original n=15, extended n=22; HUD original n=3, extended n=3), intranasal (CUD original n=4, extended n=8; HUD original n=6, extended n=11), intravenous (CUD extended n=1; HUD original n=11, extended n=17), oral (HUD extended n=1), and smoking and intranasal (original CUD n=1, extended CUD n=1).

Exclusion criteria were: 1) present or past history of DSM-IV or DSM-5 diagnoses of psychotic disorder (e.g., schizophrenia) or neurodevelopmental disorder (e.g., autism); 2) history of head trauma with loss of consciousness (> 30 min); 3) history of neurological disorders including seizures; 4) current use of any medication (with the exception of medication assisted treatment in the HUD) that may affect neurological functions; 5) current evident infection and/or medical illness including cardiovascular disease (e.g., high blood pressure), as well as metabolic, endocrinological, oncological or autoimmune diseases, and infectious diseases common in individuals with SUD including Hepatitis B and C or HIV/AIDS for the HUD group; 6) MRI contraindications including any metallic implants, pacemaker device, or pregnancy. We did not exclude SUD subjects for history of other drug addiction (e.g., alcohol, marijuana, stimulants/opiates) or other psychiatric disorders at high co-morbidity with drug addiction (e.g., depression, post-traumatic stress disorder); and 7) HC participants were excluded for a positive breathalyzer test for alcohol or positive urine screen for any psychoactive drugs.

### MRI data acquisition

MRI scans were acquired using a Siemens 3.0 Tesla Skyra (Siemens Healthcare, Erlangen, Germany) with a 32-channel head coil. T1-weighted anatomical image acquisition parameters were as follows: 3D MPRAGE sequence with 256 × 256 × 179 mm^3^ FOV, 0.8 mm isotropic resolution, TR/TE/TI=2400/2.07/1000 ms, 8° flip angle with binomial (1, −1) fat saturation, 240 Hz/pixel bandwidth, 7.6 ms echo spacing, and in-plane acceleration (GRAPPA) factor of 2, with a total acquisition time of approximately 7 min.

### Voxel-based morphometry

We followed the optimized VBM approach as documented by Good *et al*. (46) using *FSL-VBM* (47,48). First, raw DICOM images were converted to the NIFTI format via *dcm2niix* (49) and adapted to Brain Imaging Data Structure (BIDS) standards to enhance portability and reproducibility (50). Structural images were skull-stripped using FSL’s brain extraction tools and segmented into gray matter, white matter, and cerebrospinal fluid using FSL’s *fast*. FSL’s *sienax* was used to calculate normalized total brain volume (TBV). Next, all skull-stripped images were nonlinearly registered to the standard gray matter ICBM-152 template and averaged to yield an unbiased (inclusive of all participants, equally representing each group) study-specific, isotropic 2 mm template. Individual gray matter images were re-registered to this study specific template, modulated to account for registration-related warping, and spatially smoothed using a Gaussian kernel (7 mm full-width at half maximum).

To address our *a priori* hypotheses, we employed the general linear model approach using our original sample (n=60). We included group (HC, CUD, and HUD), age, and total brain volume (TBV) as regressors in the model to produce age- and TBV-corrected linear contrasts comparing HC to both SUD groups (an “SUD” map encompassing both CUD and HUD), HC to CUD and HUD separately, and the CUD and HUD groups to each other. We accounted for the potential contribution of other explanatory variables on our results by first comparing HC, CUD, and HUD groups on demographic and neuropsychological measures using one-way ANOVAs (and pairwise comparisons where appropriate) for continuous and chi-square tests (or Fisher’s exact tests where appropriate) for categorical variables, corrected for familywise error (α=.05/8=.006 for the eight comparisons between HC, CUD, and HUD; α=.05/6 for the six comparisons between CUD and HUD). Then, variables showing significant group differences were entered into VBM correlations to detect potential relationships with GMV across all participants. Those with significant group differences and GMV correlations were used as controlled variables in VBM analyses to correct for their potential contribution to the group differences in GMV.

For SUD group comparisons in drug use-related patterns as a function of GMV, we used separate models for each regressor (the use variables that did not differ by group: days since last use, lifetime use in years, craving, withdrawal, Table 1) to avoid multicollinearity, and compared the slopes of the correlations between CUD and HUD groups to derive CUD- and HUD-specific correlation maps (corrected for familywise error, *p*<.05/4=.012). For completeness, we also inspected severity of dependence in VBM analyses independently from the above. We did not examine past month use, as only four out of the 32 individuals with HUD in the extended sample reported non-zero values. To look for potential group similarities, we also tested correlations between these drug use measures with GMV using the whole-brain analyses in the combined SUD group (both samples).

Because parametric methods rely on a Gaussian distribution assumption that is difficult to achieve due to the inherent noise in MRI data (51), unlike prior inspections of GMV differences between CUD and HUD (36), we used permutation-based non-parametric statistical tests using FSL’s *randomise* to compute all contrasts. We applied 5,000 permutations per test and threshold free cluster enhancement (corrected to *p*<.05 for primary group differences and *p*<.012 for the four drug use variable models) to detect significant clusters (52). We repeated these VBM analyses using our larger replication sample (n=96), reporting small volume corrected results within an *a priori* selected cortico-striatal anatomical mask. This combined mask encompassed the vmPFC (inclusive of frontal medial, frontal orbital, and subcallosal cortices), striatum (inclusive of NAcc, caudate, putamen), and IFG (inclusive of pars opercularis and triangularis subregions), derived from the Harvard-Oxford Cortical/Subcortical Atlas with a 50% probabilistic threshold applied to each region.

## Results

### Participants

Groups were comparable in age, sex, education, verbal and non-verbal IQ in both the original and extended samples (*p*≥.121; see Table 1). Group differences were evident in race in both samples (driven by fewer Black and more White participants in the HUD group compared to the other groups) and self-reported depression in the extended sample (driven by higher scores in HUD compared to HC). However, race and depression did not display a significant relationship with GMV across the original or the replication samples (all *p*s≥.159), and thus were not included as controlled variables in testing our main hypotheses. The three groups further differed in cigarette smoking status (fewest current smokers in the HC group); however, there were no significant differences in smoking status between HUD and CUD groups [original: Fisher’s exact test *p*=.106; extended: Fisher’s exact test *p*=.011, FWE-corrected]. Finally, past month use was by definition lower in the inpatient HUD than CUD in both the original (at a trend level) and extended samples. However, despite trends for lower lifetime use in the HUD than CUD group, the former group showed significantly higher severity of dependence in both samples.

### SUD compared to HC

Compared to HC, the SUD group showed significantly lower GMV in the vmPFC (*p*=.039; Table 2), as also reflected by a trend in the replication sample (*p*=.059).

**Table 2.** Substance-general and specific gray matter volume differences.

|  | Original sample (N=60) |  |  |  |  |  | Replication sample (N=96) |  |  |  |  |  |
| --- | --- | --- | --- | --- | --- | --- | --- | --- | --- | --- | --- | --- |
|  |  |  |  | MNI |  |  |  |  |  | MNI |  |  |
| Contrast | Region | Voxels | *p* | x | y | z | Region | Voxels | *p* | x | y | z |
| HC>SUD | Ventromedial PFC | 102 | .039 | -4 | 36 | -26 | Ventromedial PFC <sup>†</sup> | 38 | .059 | -4 | 30 | -28 |
| SUD>HC | No significant clusters |  |  |  |  |  | No significant clusters |  |  |  |  |  |
| HC>CUD | No significant clusters |  |  |  |  |  | No significant clusters |  |  |  |  |  |
| CUD>HC | No significant clusters |  |  |  |  |  | No significant clusters |  |  |  |  |  |
| HC>HUD | Ventromedial PFC | 325 | .013 | 2 | 26 | -24 | Ventromedial PFC | 167 | .006 | -4 | 28 | -26 |
|  | Nucleus accumbens | 69 | .045 | -10 | 12 | -10 | Putamen/Nucleus accumbens | 86 | .037 | -16 | 8 | -12 |
|  | Orbitofrontal cortex | 3 | .049 | 12 | 6 | -22 | Orbitofrontal cortex <sup>†</sup> | 8 | .091 | 14 | 8 | -18 |
|  | Orbitofrontal cortex | 50 | .042 | -14 | 6 | -24 | Caudate <sup>†</sup> | 22 | .082 | 14 | 4 | 12 |
|  | Pons | 1,801 | .004 | 12 | -18 | -38 | -- | -- | -- | -- | -- | -- |
| HUD>HC | No significant clusters |  |  |  |  |  | Inferior frontal gyrus | 166 | .010 | -56 | 16 | 16 |
| HUD>CUD | Inferior frontal gyrus | 129 | .029 | 56 | 18 | 6 | Inferior frontal gyrus | 347 | .005 | 56 | 18 | 8 |
| CUD>HUD | Pons | 257 | .033 | 8 | -14 | -34 | -- | -- | -- | -- | -- | -- |
|  | -- | -- | -- | -- | -- | -- | Ventromedial PFC <sup>†</sup> | 11 | .074 | -4 | 22 | -26 |

Clusters are corrected to $p < .05$ following permutation-based non-parametric hypothesis testing and threshold-free cluster enhancement. <sup>†</sup>: Clusters reflecting trends in replication analyses as identified by one-tailed t-tests. Voxel locations are presented in MNI space, sorted by y-axis coordinates (anterior to posterior).

### CUD compared to HC

There were no significant results when comparing the CUD group to the HC group in either the main whole-brain or the small volume corrected replication VBM analyses.

### HUD compared to HC

Compared to the HC, the HUD group showed significantly lower GMV in the vmPFC (*p*=.013), left NAcc (*p*=.045), bilateral OFC (left: *p*=.042, right: *p*=.049), and pons (*p*=.004) (Figures 1-2, Table 2). The replication analysis also revealed significant GMV reductions in the HUD group compared to HC in the vmPFC (*p*=.006) and the left NAcc (*p*=.037; Figures 1-2, Table 2), with trends in the right OFC (*p*=.091) and caudate (*p*=.082). Note that the pons was not a region of *a priori* interest, and was not included in our replication mask for small volume correction. While the original sample did not reveal significantly higher GMV in HUD compared to HC, the small volume corrected replication indicated higher left IFG GMV in HUD (*p*=.010).

**Figure 1.**
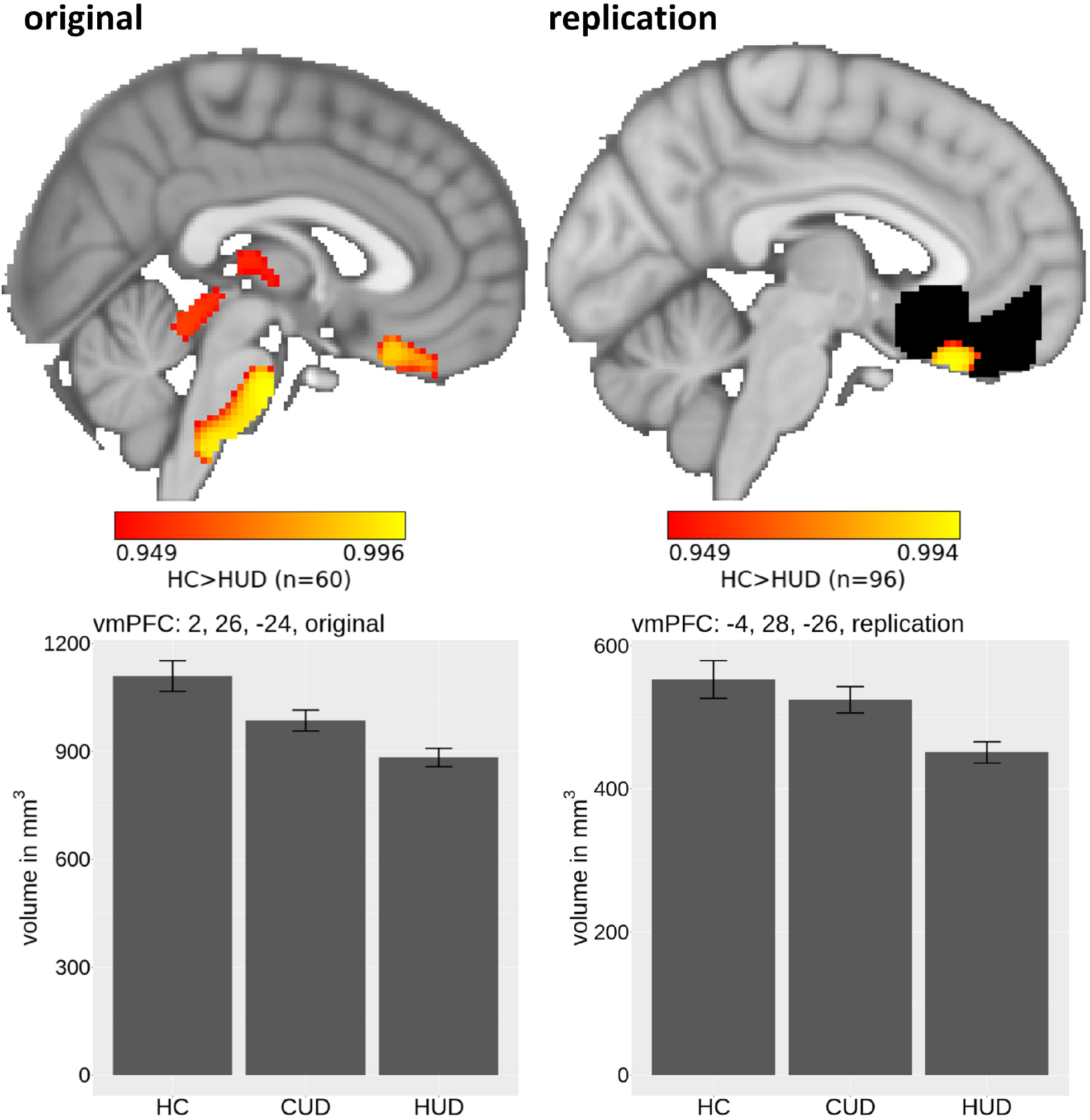
Ventromedial prefrontal cortex gray matter volume reductions in heroin use disorder. Heroin-addicted individuals in the original sample (n=60) exhibited significantly reduced ventromedial prefrontal cortex gray matter volume (left panel; whole-brain corrected to *p*<.05) compared to healthy control subjects. This effect showed a similar trend (*p*=.059) in the extended sample (n=96; right panel) within an *a priori* combined bilateral mask of the ventromedial prefrontal cortex, inferior frontal gyrus, and striatum denoted in white (small volume corrected to *p*<.05). Contrasts in both analyses were corrected for age and total brain volume.

**Figure 2.**
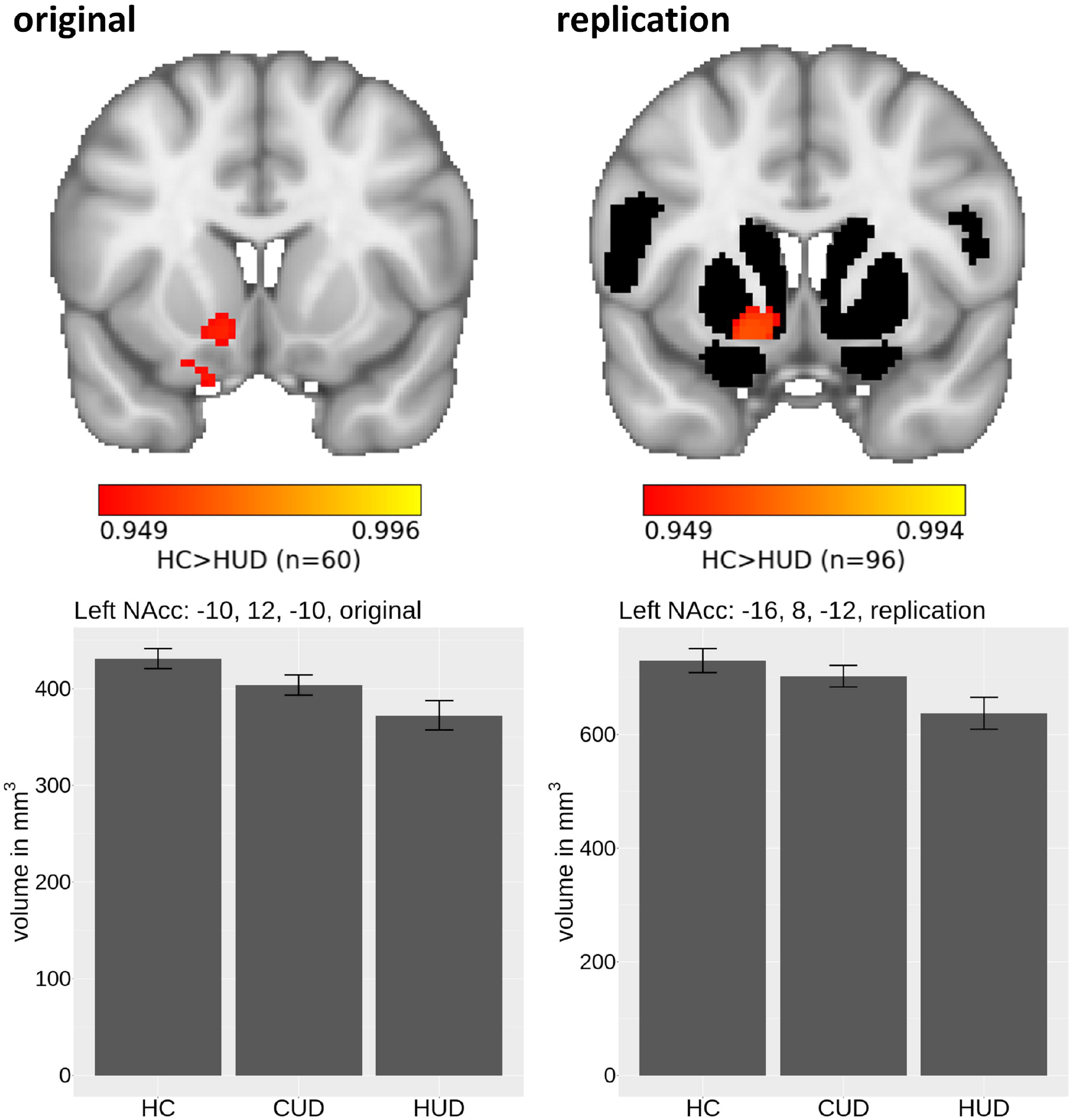
Nucleus accumbens gray matter volume reductions in heroin use disorder. Heroin-addicted individuals in the original sample (n=60) exhibited significantly reduced left nucleus accumbens gray matter volume (left panel; whole-brain corrected to *p*<.05) compared to healthy control participants. This effect was replicated in the extended sample (n=96; right panel) within an *a priori* combined bilateral mask of the ventromedial prefrontal cortex, inferior frontal gyrus, and striatum denoted in white (small volume corrected to *p*<.05). Contrasts in both analyses were corrected for age and total brain volume.

### CUD compared to HUD

The direct contrast between CUD and HUD indicated that the former group showed significantly lower right IFG GMV (*p*=.029) (Figure 3, Table 2). This effect persisted in the replication analysis (right: *p*=.005).

**Figure 3.**
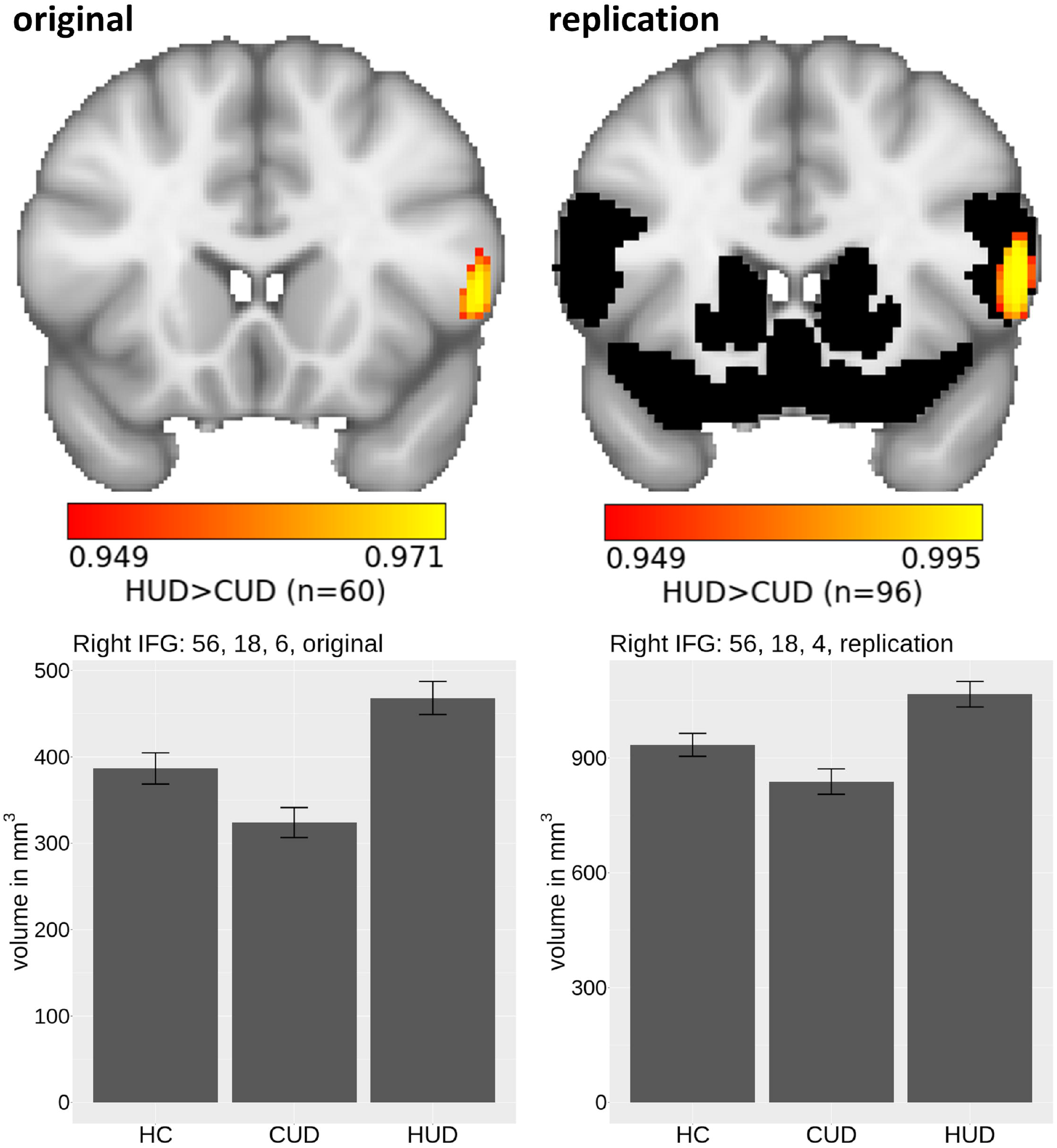
Inferior frontal gyrus gray matter volume reductions in cocaine compared to heroin use disorder. Cocaine-addicted individuals in the original sample (n=60) exhibited significantly reduced right inferior frontal gray matter volume (left panel; whole-brain corrected to *p*<.05) compared to heroin-addicted individuals. This effects was replicated in the extended sample (n=96; right panel) within an *a priori* combined bilateral mask of the ventromedial prefrontal cortex, inferior frontal gyrus, and striatum, denoted in white (small volume corrected to *p*<.05). Contrasts in both analyses were corrected for age and total brain volume.

The CUD>HUD contrast revealed significantly higher pons GMV in CUD (*p*=.033). We did not test this effect in the replication analysis. However, a trend was evident in the vmPFC GMV (CUD>HUD, *p*=.074).

### Whole-brain GMV and addiction severity correlations

Whole-brain GMV correlations with the drug use variables revealed that the higher the posterior putamen GMV, the higher the craving in the CUD compared to the HUD group (Figure 4, α=.012; 111 voxels, peak MNI coordinates 34, -8, 8, *p*=.011). This correlation was supported (α=.012) in the replication analysis (19 voxels, peak MNI coordinates 32, -8, 8, *p*=.039, small volume corrected). No other drug use variable correlated significantly with GMV using the whole-brain analyses in CUD or HUD or the combined SUD groups, in the original or extended samples.

**Figure 4.**
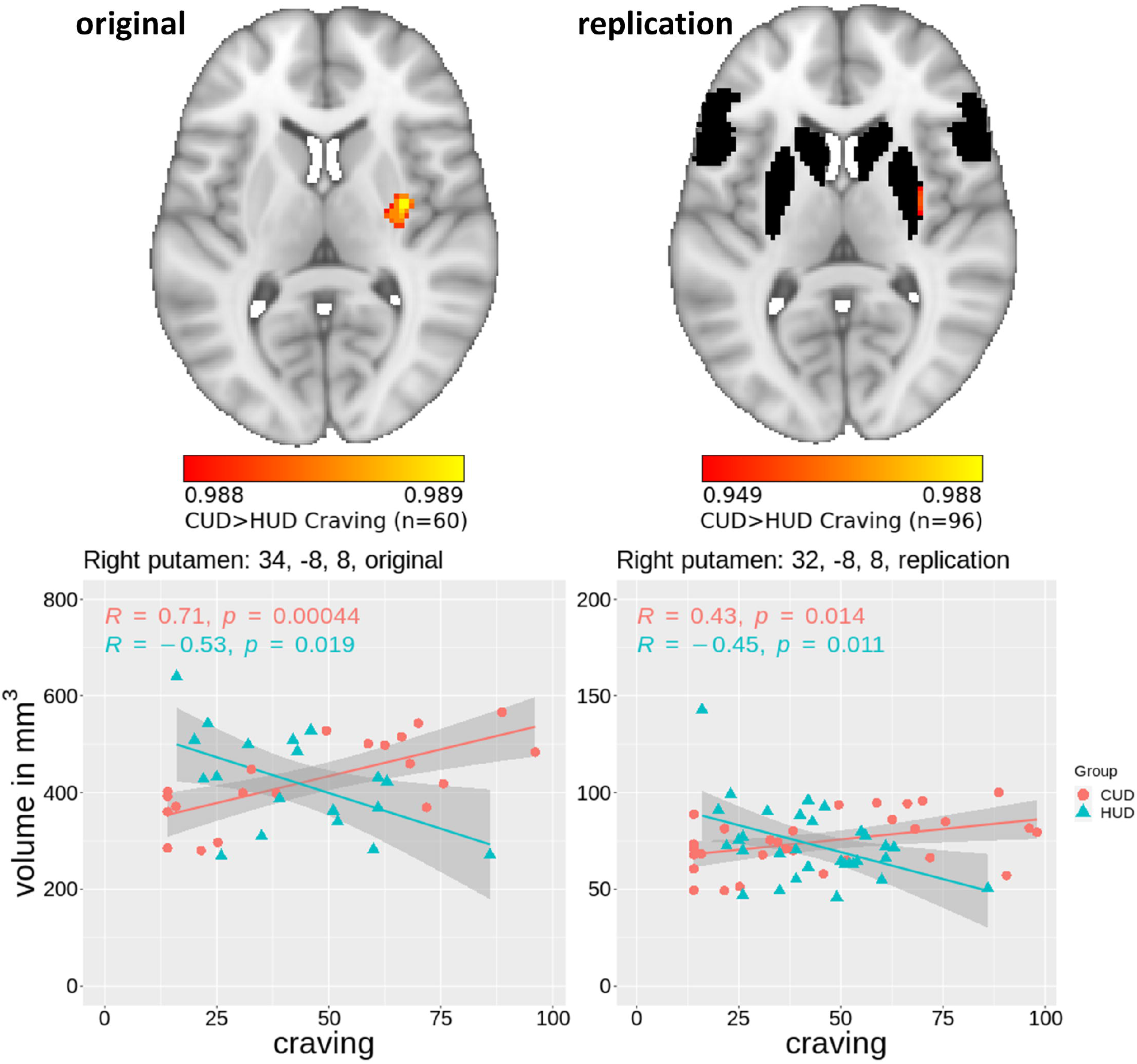
Increased posterior putamen volume as a function of drug craving in cocaine compared to heroin use disorder. Posterior putamen volume was more positively correlated with cocaine craving in cocaine-addicted individuals compared to heroin craving in heroin-addicted individuals in our original sample (n=60; left panel; whole-brain corrected to *p*<.012 for multiple comparisons). This effect was supported in the extended sample (n=96; right panel) within an *a priori* combined bilateral mask of the ventromedial prefrontal cortex, inferior frontal gyrus, and striatum denoted in white (small volume corrected, *p*=.039). Contrasts in both analyses were corrected for age and total brain volume.

## Discussion

Although GMV reductions in individuals with SUD have been commonly documented (53), research directly interrogating the common and distinct patterns in human CUD and HUD has been limited, largely highlighting posterior (temporo-parietal and cerebellar) volumetric differences between these different drug types (36,37). Here, using demographically well-matched groups of CUD, HUD, and HC, we identified cortico-striatal similarities and differences in GMV changes (from HC) between these drug classes and internally replicated our results in an extended sample of participants. First, we replicated the previously reported reductions in vmPFC GMV in addicted individuals in our original sample, showing a similar trend in the replication sample, as driven by the HUD group. Second, the HUD group also showed lower NAcc GMV, discernable when compared to the HC group. Third, when directly compared to the HUD group, the CUD group showed significantly lower right IFG GMV and, fourth, this direct contrast also revealed a unique (positive) correlation in the CUD group between drug craving and the posterior putamen.

In both SUD groups, and driven by the HUD, lower vmPFC GMV compared to HC replicates similar findings in addicted individuals across substance types as reported by studies using meta-/mega-analyses (6,7), and those focused on stimulants (34,54–59) and opiates (35,60,61) independently. Drug use may play a causal role in structural vmPFC degradation (53) as suggested by decreased vmPFC volume following randomization to chronic cocaine self-administration in non-human primates (62), and this region’s volumetric recovery with abstinence from cocaine (as associated with recovery of relevant neuropsychological functions including decision-making) in humans (63). Alternatively, alterations in vmPFC gray matter may be a biomarker of addiction vulnerability, as lower vmPFC/OFC cortical thickness predicts increased adolescent drug experimentation (number of drugs tried) in individuals exposed to maternal cigarette smoking during gestation (64). The HUD group also showed NAcc decreases, a result that extends prior efforts in humans (26) and is consistent with evidence of decreased NAcc neuroplasticity following chronic opiate self-administration in rodents (21). Lack of such an effect in the CUD is consistent with the mixed prior evidence in chronic psychostimulant use (65), with studies showing both higher (66) and lower NAcc volume compared to HC (67).

Several mechanisms may underlie these drug associated volumetric reductions in both the vmPFC and NAcc encompassing reduced tonic dopamine neurotransmission mediated by striatal medium spiny neurons that serve as a dopaminergic input nexus from the PFC (68,69). Indeed, dopamine receptor D2/3 availability (measured via PET with [^18^F] Fallypride and [^11^C] Raclopride) is reduced in human drug addiction (70,71)] as associated with decreased gray matter (evaluated with VBM) in the medial PFC and NAcc (4). Other mechanisms affecting the PFC, and especially the vmPFC/anterior PFC, include cerebral glucose metabolism reductions following chronic psychostimulant use as documented using 2-[^14^C] Deoxyglucose in non-human primates (72,73) and [^18^F] Fluorodeoxyglucose in humans (74–76). The severity of deficit in the HUD group is possibly also driven by cellular downregulation [measured by the phosphorylation of protein kinases such as the ERK and MAPK that play a role in neuronal growth (77) and are activated by neurotrophic factors (78)], as suggested by post-mortem evaluations of PFC cytoarchitecture in opiate-addicted individuals (heroin or methadone) (79). Interestingly, ERK phosphorylation in the NAcc is also decreased following chronic (but not acute) morphine administration (80). Results in humans in vivo are consistent with these suggestions as documented by decreased measures of neuronal integrity in mostly medial PFC gray matter assessed with magnetic resonance spectroscopy (via *N*-acetyl compounds and myoinositol) in chronic users of heroin (81) and cocaine (82). Taken together, these results of reduced mesocorticolimbic integrity especially in individuals with HUD need to be explored vis-à-vis functional measures of salience/value processing, extinction learning, and goal-directed control.

The volumetric right IFG compression in the CUD compared to the HUD group in our study agrees with extensive evidence for lower IFG GMV in chronic stimulant users compared to HC (34,57,86,87). Given the well-established role of the right IFG in impulsivity and inhibitory control (29–31,88), and the heightened impulsivity characterizing psychostimulant use disorder [cocaine (5,32,57) and methamphetamine (87)], these results suggest a potential correlate in CUD for deficits in impulse control. Accordingly, we also found a CUD-specific positive correlation between the posterior putamen GMV and drug craving. The posterior putamen is a node of the habit network (89,90), and activity in this region increases as a function of stimulus-response training reflective of sensitivity to salient cues (17), which, in CUD, may be related to the drug cue-induced dorsal striatal dopamine release as associated with craving (18). Posterior putamen gray matter density negatively correlates with behavioral indices of goal-directed control, such that the higher its density, the worse (more habitual) the performance (91), as potentially mediated by reduced glutamate concentration and turnover in CUD (92). Taken together, the combined IFG and putamen GMV profiles in the current study suggest a unique marker towards an enhanced propensity to develop impulsive drug-seeking behaviors specifically in the CUD.

There were several null and/or unexpected results in this study. First, although previously reported (53,54), we did not find a significant relationship between decreased vmPFC/OFC GMV and longer duration of drug (including cocaine) use. Second, contrary to prior evidence (35), the HUD in the extended (but not original) sample exhibited higher left IFG GMV than HC. Third, while putamen GMV is generally reported to be increased in drug addiction (15) [although decreases in this region were also reported in chronic opioid users (93)] we did not observe putamen GMV differences between our groups. Future efforts to resolve these discrepancies may benefit from larger samples that should also be more closely matched in drug use patterns such as recent use frequency and dependence severity. Furthermore, considering that abstinence from drug use has been associated with cortico-striatal recovery in addicted individuals (94), the longitudinal inspection of treatment-related effects is needed. Relatedly, since tobacco use has been linked to decreases in PFC GMV (33,95), accounting for smoking-related patterns in more balanced samples (e.g., including more HC participants who smoke cigarettes) is warranted. Sex-specific IFG GMV compression has been documented in cocaine-addicted individuals such that cocaine-addicted women exhibit lower IFG [and insula (96)] GMV than non-addicted women (97), warranting a closer inspection of potential sex differences and hormonal effects in substance-specific neural morphology. The CUD and HUD groups further differed in race; however, race did not yield a significant relationship with GMV in our results, in line with the lack of race-related gray matter concentration patterns in CUD (55). Finally, while we report key differences in the vmPFC, NAcc, IFG, and putamen morphology, behavioral measures (e.g., of drug cue reactivity and its extinction, value based decision-making, reward prediction error, impulsivity, and habit formation) would be needed to link these anatomical results to function.

To the best of our knowledge, this is the first demonstration of both unique and overlapping prefrontal cortico-striatal gray matter morphology changes in human cocaine and heroin addiction, providing insight into the brain-related variability accompanying the use of these different drug classes. Specifically, we demonstrate vmPFC GMV compression across both CUD and HUD combined (with a more severe pattern in the latter) in addition to NAcc decreases in HUD, and IFG reductions in CUD. Together with the IFG compression, the unique relationship between increased cocaine craving and increased posterior putamen GMV underscores a potential biomarker in CUD, suggestive of a disease profile that may be conducive to cue-triggered habits/impulsive behaviors. In general, these results extend patterns observed in rodent models of chronic drug self-administration to addiction in humans, alluding to the cross-species conservation of cortico-striatal alterations with the chronic use of psychostimulants and opiates. These results call for closer examination of differences between drug classes in functions supported by these brain regions inclusive of select behaviors, cognitions and emotions and functional network organizations. Importantly, these findings underscore the importance of treating the human addiction experience as a multifaceted disease that takes into account the primary drug of abuse and warrants tailored interventions.

## Data Availability

All data produced in the present study are available upon reasonable request to the authors.

## Acknowledgments

This work was supported by 1R01AT010627 and 1R01DA048301 to RZG. This manuscript has been posted on a preprint server.

## Disclosures

Ahmet O. Ceceli declares no conflicts of interests. Yuefeng Huang declares no conflicts of interests. Greg Kronberg declares no conflicts of interests. Pias Malaker declares no conflicts of interests. Pazia Miller declares no conflicts of interests. Sarah King declares no conflicts of interests. Pierre-Oliver Gaudreault declares no conflicts of interests. Natalie McClain declares no conflicts of interests. Lily Gabay declares no conflicts of interests. Devarshi Vasa declares no conflicts of interests. Defne Ekin declares no conflicts of interests. Nelly Alia-Klein declares no conflicts of interests. Rita Z. Goldstein declares no conflicts of interests.

## Notes

### Competing Interest Statement

The authors have declared no competing interest.

### Author Declarations

The Icahn School of Medicine at Mount Sinai institutional review board approved study procedures and all participants provided written informed consent.

